# Evaluating the Relationship between Psychological Traits and Resilience to Musculoskeletal Injuries in Combat Control Graduates by Mendelian Randomization

**DOI:** 10.1101/2023.05.16.23289997

**Authors:** Richard R. Chapleau

## Abstract

U.S. Air Force combat control (CCT) personnel are a group of highly trained personnel performing a wide range of aviation-related tasks in contested combat environments. Certified by the U.S. Federal Aviation Administration to conduct air traffic control operations, CCTs are required to maintain high levels of alertness and perform complex tasks in high stress, high threat environments. Those CCTs who complete training are in exceptional physical, psychological, and cognitive fitness, however nearly 70% of CCT candidates will develop some form of musculoskeletal injury (MSI) during training. Using only open-source summary statistics results of genome-wide association studies (GWAS) on civilian populations, we report our findings from two-sample Mendelian randomization (MR) estimates evaluating the causal relationships between personality and psychological strengths associated with success in the CCT training program and MSI. We used the TwoSampleMR R-package and GWAS statistics obtained from the IEU OpenGWAS project with instrumental variables extracted at GWAS-significant and suggestive thresholds (*P* < 5×10^−8^ and 5×10^−5^, respectively). Back pain and dislocations were the most common outcomes caused by personality and psychological traits. Altogether more than 150 MSI outcomes were identified with causes related to psychological traits associated with successfully completing combat control training. The implications of our results suggest that the combat control training program, and by extension other special operations training programs, could encounter fewer injuries by encouraging utilization of embedded psychological assets.

## Introduction

In the US military, the combat control (CCT) training pipeline has a historical attrition rate between 70 and 80 percent [1], nearly double the programmed or expected attrition rates [2]. The majority of the washback and attrition occurs in the orientation and apprentice courses [2]. The primary factor in attrition is self-elimination (approximately 70%), followed by medical (14%) and physical issues (12%). Many of the self-eliminations are caused by injuries or insufficient fitness (22% of trainees self-eliminating reported the reason as physical). Interestingly, only a small fraction (<2%) of the eliminations occurred because of discipline or mental health (e.g., hydrophobia) issues, and they accounted for approximately 10% of self-eliminations.

Those trainees who ultimately become CCT graduates are mentally tough, scoring 7.9 out of 10 on the Mental Toughness Questionnaire [1]. They are confident, resilient, persistent, and positive when facing challenges. They also have high extraversion and conscientiousness, and low neuroticism and openness to experience [1]. These personality traits help them work well in small and dynamic teams. CCT graduates have higher cognitive aptitude than non-graduates, especially in arithmetic and math [3]. They also have higher emotional intelligence than non-graduates and the general population [3]. They can cope with stress, self-regulate, and empathize. These aptitudes in math and emotional skills may increase their chances of completing the CCT training program successfully. Therefore, we suspect that the selection process chooses recruits with the psychological stamina and resilience to absorb the demanding 175 days of technical training.

The psychological state of the athlete has been associated with preventing injury and promoting recovery through qualitative assessments from coaches, athletes, and sports medicine physicians [4-6]. Additionally, quantitative assessments reinforced these qualitative findings [7-18]. Anxiety and similar traits like worry and concern over mistakes have been repeatedly observed for a wide range of athletic activities [7,8,11-15]. Furthermore, psychological traits also can increase resilience to injury, such as having a high perception of one’s physical energy and health [19], being self-confident [19], practicing positive coping strategies [9], having a mastery climate [16], and having a sense of hardiness or resiliency [18]. The literature suggests a significant relationship between psychology and injury, but to our knowledge, no randomized trials have been performed to evaluate causal directionality, likely due to the infrequent and seemingly random nature of MSI.

Therefore, we hypothesized that the psychological strengths of CCT graduates made them less likely to get physical injuries that could stop them from completing the training pipeline. We used Mendelian randomization (MR) [20] to test our hypothesis, which is a statistical method that uses genetic variants correlated with an exposure trait (e.g., smoking) but not the outcome (e.g., disease) to see if the trait causes the outcome. Here, we used MR to see if psychological traits linked to CCT graduation (the exposures) caused musculoskeletal injuries (the outcomes). MR has been used before to study factors that cause MSI [21, 22], but to the best of our knowledge it has not been used to assess the psychological factors causing injury.

## Methods

### Psychological trait identification

A literature search was performed to identify psychological traits relating to successful completion of the training for combat control (CCT). Specifically, we searched for publicly accessible papers using PubMed. The search criteria used was “combat control” or “special tactics” and “graduation” or “training” and “psychological” or “selection” or “behaviors.” These journal articles were supplemented with limited access studies published in the Defense Technical Information Center (DTIC).

We included all studies that reported psychological traits associated with the outcome of interest, whether quantitative or qualitative in design. We excluded studies that reported solely cognitive (e.g., scores from the Armed Services Vocational Aptitude Battery) or physical (e.g., 1.5 mile run time) traits. Since CCT is a unique career field within the U.S. military, we excluded studies performed in non-U.S. service-members. We included all traits reported from the studies regardless of if additional studies support or rebut their inclusion. Additionally, we considered CCT graduation and emotional reactions to killing independently, meaning that if a trait was related to both, then we included it in both lists.

Explicit inclusion criteria for studies related to completing CCT training:

1. Study must be primary literature (reviews, books, meta-analyses, editorials, commentaries and similar were excluded);
2. Participants were US military service members who had entered CCT training (other special warfare career fields and non-Air Force special operations forces were excluded as this study’s focus is on the CCT operator);
3. Participants completed a personality inventory or emotional intelligence questionnaire;
4. Psychological traits identified were reported alongside the inventory or questionnaire used; and
5. Participants were classified based upon completing the full training pipeline or attrition due to either self-elimination or cadre-determined failure.

### GWAS identification

We searched the IEU OpenGWAS Project [23] for each psychological trait identified and for traumatic musculoskeletal injuries (fractures, tears, sprains, strains, and dislocations), pain, and joint damage (bursitis, tendinitis, synovitis, etc.). We restricted fractures to exclude those phenotypes caused by osteoporosis; involving operative procedures; occurring to the face, skull, or head; or resulting from specific causes (e.g., “falls resulting from simple fall”). We similarly restricted pain to those phenotypes only located in joints, limbs, or muscles (e.g., no generalized, chest or abdominal pain) and excluded phenotypes involving treatments or pain medications. Finally, we did not include sub-populations studies since not all phenotypes were divided by populations. For example, the phenotype “knee pain for 3+ months” included the two studies “ukb-e-3773_CSA” and “ukb-b-8906”, and we excluded the former. The OpenGWAS Project contains the summary statistic (aggregate data) results from 40,027 GWAS datasets (as of April 2022) and uses a standardized format for storing the data [24]. The MSI phenotypes used are provided in Table 1 and study identifiers are included in Supplementary Table 1.

**Table 1:** Musculoskeletal injury phenotypes used in this study.

| **Musculoskeletal Injury OpenGWAS Phenotypes** |  |
| --- | --- |
| Achilles tendinitis | Fractured heel (left) |
| Back pain for 3+ months | Fractured heel (right) |
| Diagnoses - main ICD10: M23.2 Derangement of meniscus due to old tear or injury | Fractured/broken bones in last 5 years |
| Diagnoses - main ICD10: M23.22 Derangement of meniscus due to old tear or injury (Posterior cruciate ligament or Posterior horn of medial meniscus) | Diagnoses - main ICD10: M23.23 Derangement of meniscus due to old tear or injury (Medial collateral ligament or Other and unspecified medial meniscus) |
| Hip pain for 3+ months | Low back pain |
| Diagnoses - main ICD10: M25.5 Pain in joint | Lower back pain or/and sciatica |
| Diagnoses - main ICD10: M54.5 Low back pain | Muscle strain |
| Diagnoses - main ICD10: M54.56 Low back pain (Lumbar region) | Non-cancer illness code, self-reported: back pain |
| Diagnoses - main ICD10: M54.59 Low back pain (Site unspecified) | Non-cancer illness code, self-reported: fracture lower leg / ankle |
| Diagnoses - main ICD10: M65 Synovitis and tenosynovitis | Diagnoses - main ICD10: S52.50 Fracture of lower end of radius (closed) |
| Diagnoses - main ICD10: S52 Fracture of forearm | Other/unspecified synovitis and tenosynovitis |
| Non-cancer illness code, self-reported: joint pain | Pain in joint |
| Dislocation, sprain and strain of joints and ligaments at ankle and foot level | Dislocation, sprain and strain of joints and ligaments of elbow |
| Pain in limb | Pain type(s) experienced in last month: Back pain |
| Dislocation, sprain and strain of joints and ligaments of knee | Pain type(s) experienced in last month: Hip pain |
| Fracture of foot, except ankle | Pain type(s) experienced in last month: Knee pain |
| Fracture of forearm | Pathological/recurrent dislocation and subluxation of joint, not elsewhere classified |
| Fractured bone site(s): Ankle | Peroneal tendinitis |
| Fractured bone site(s): Arm | Recurrent dislocation and subluxation of joint |
| Fractured bone site(s): Other bones | Transient synovitis |
| Fractured heel |  |

### Mendelian randomization

We performed two-sample Mendelian randomization (2SMR) estimates using the TwoSampleMR R package [25]. We used the MR Egger regression [26], inverse variance weighted (IVW) estimator [27], weighted median estimator [28], and Wald ratio estimator [29] algorithms. Instrumental variables (IV, SNPs associated with the exposure) were extracted by *P*-value thresholds 5×10^−5^ and 5×10^−8^. We excluded SNPs in strong linkage disequilibrium (LD) to reduce bias and used a clumping process with European samples from the 1,000 Genomes Project (R^2^ < 0.001, window size = 10,000). If SNPs identified in the exposure dataset were not in the outcome dataset, proxy SNPs in LD (r^2^ > 0.9) were used as instrumental variables. For the sensitivity analysis, we performed heterogeneity testing using Cochran’s Q and I^2^ analyses [29] and tested pleiotropy on the weighted median estimation results [28].

## Results

### Literature review identifies psychological traits associated with graduating CCT training

Our literature search identified 5 studies reporting psychological traits related to graduating from CCT training (Figure 1). We identified 29 traits associated with graduating CCT training from the psychological literature (Table 2). These included the aforementioned associations of NEO personality inventory domains extraversion and conscientiousness [17, 31] and traits from the EQI [19]: general mood, intrapersonal, stress management, optimism, happiness, and adaptability. Of the 29 traits, 15 were found in the EQI and the remaining 14 were from the NEO personality inventory.

**Table 2:** Psychological traits associated with successfully completing CCT training.

| Graduating CCT Training |  |  |
| --- | --- | --- |
| Adaptability (EQI)<sup>a</sup> | Flexibility (EQI)<sup>a</sup> | Self-actualization (EQI)<sup>a</sup> |
| Agreeableness (NEO)<sup>b</sup> | General mood (EQI)<sup>a</sup> | Self-regard (EQI)<sup>a</sup> |
| Anxiety (NEO)<sup>b</sup> | Happiness (EQI)<sup>a</sup> | Straightforwardness (NEO)<sup>b</sup> |
| Assertiveness (EQI)<sup>a</sup> | Independence (EQI)<sup>a</sup> | Stress management (EQI)<sup>a</sup> |
| Assertiveness (NEO)<sup>b</sup> | Interpersonal relationships (EQI)<sup>a</sup> | Stress tolerance (EQI)<sup>a</sup> |
| Compliance (NEO)<sup>b</sup> | Intrapersonal (EQI)<sup>a</sup> | Tendermindedness (NEO)<sup>b</sup> |
| Conscientiousness (NEO)<sup>c</sup> | Modesty (NEO)<sup>b</sup> | Trust (NEO)<sup>b</sup> |
| Depression (NEO)<sup>b</sup> | Neuroticism (NEO)<sup>b</sup> | Values (NEO)<sup>b</sup> |
| Emotional self-awareness (EQI)<sup>a</sup> | Optimism (EQI)<sup>a</sup> | Vulnerability (NEO)<sup>b</sup> |
| Extraversion (NEO)<sup>c</sup> | Reality testing (EQI)<sup>a</sup> |  |

CCT = combat control; EQI = Emotional Questionnaire Inventory; NEO = NEO Personality Inventory; References: a) [19]; b) [30]; c) [17]

**Figure 1:**
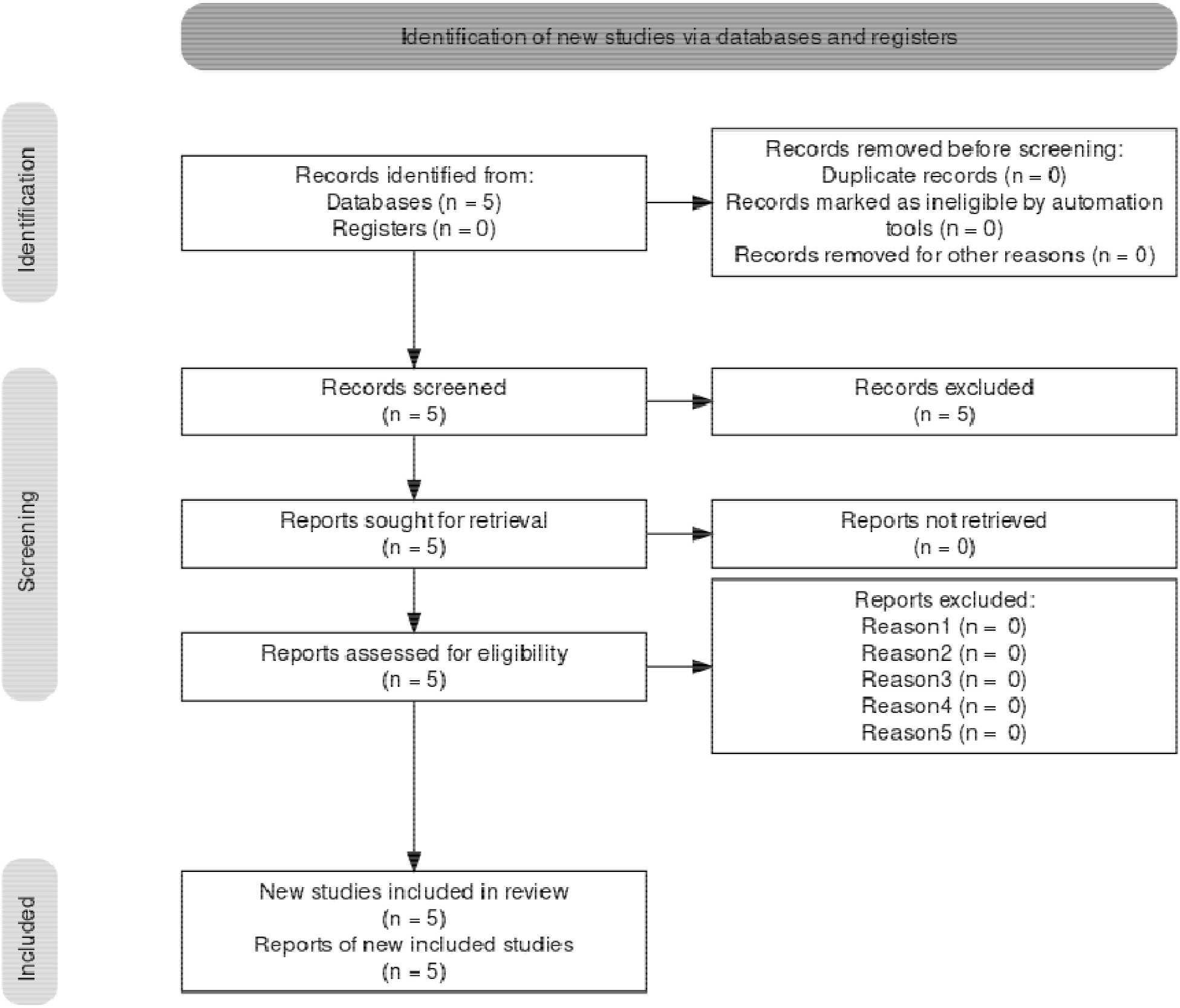
PRISMA flow diagram for literature review to identify psychological strengths of CCT graduates

From the IEU OpenGWAS project, we identified GWAS phenotypes corresponding to the aforementioned psychological traits for completing CCT training. We identified 31 distinct GWAS phenotypes for traits associated with the former (Table 3). We found only 14 traits with GWAS summary statistic datasets, but a total of 46 studies were identified. Nine traits were reported in multiple studies: neuroticism (14 studies); health satisfaction (5); seen a general practice doctor for nerves, anxiety, tension or depression (5); self-injury such as self-cutting, scratching or hitting (4); seen a psychiatrist for nerves, anxiety, tension or depression (4); anxiety/panic attacks (3); financial situation satisfaction (2), mental health problems diagnosed by a professional for anxiety, nerves, or generalized anxiety disorder (2), and a derived neuroticism score (2).

**Table 3:**
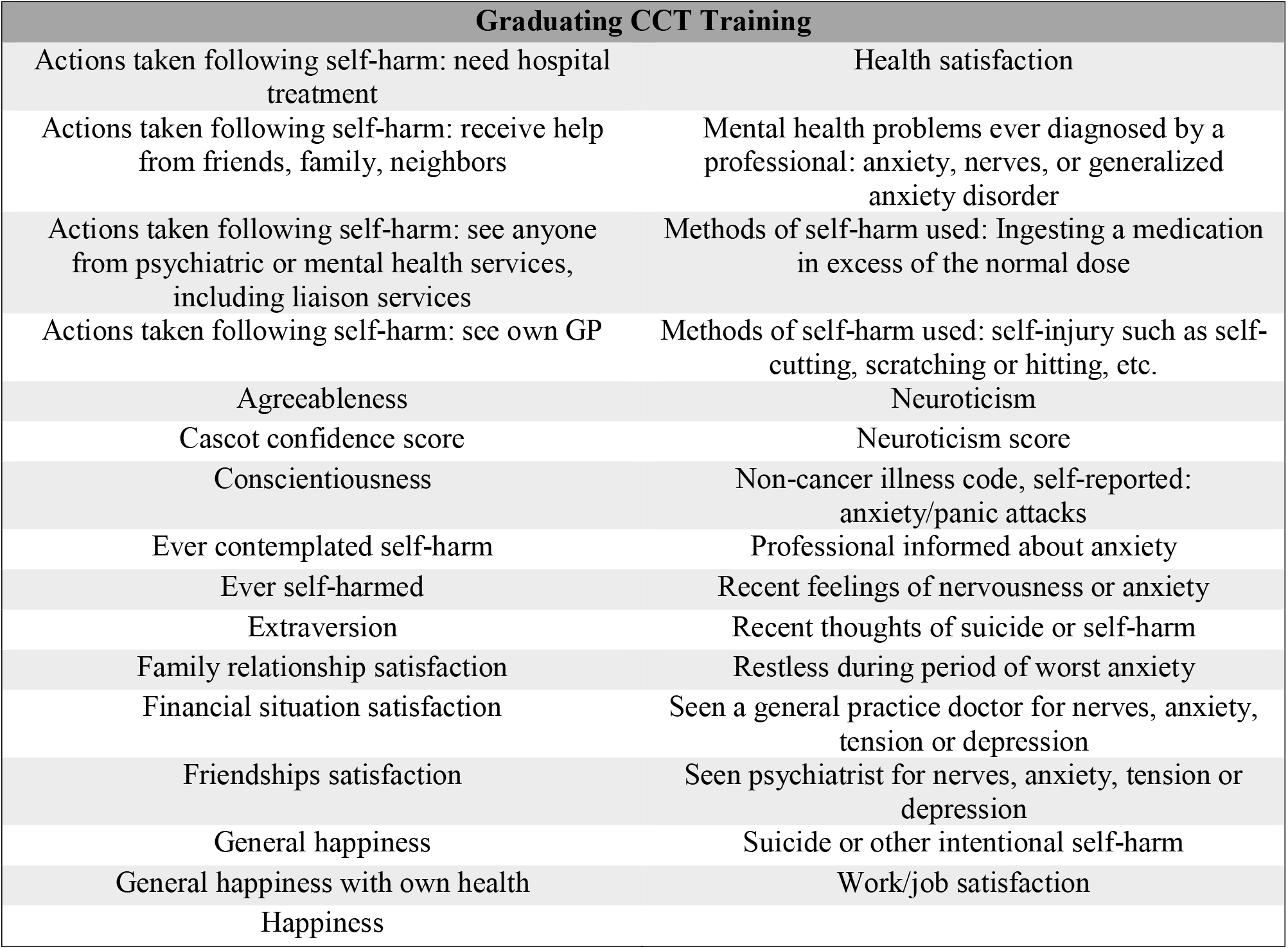
Phenotypes identified from the IEU OpenGWAS Project that are related to the traits associated with successfully completing CCT training.

### The causal influence of the NEO “Big 5” personality domains on resilience to injury

While 13 of the traits associated with successfully completing CCT training were identified from the NEO personality inventory and there were 26 GWAS datasets identified related to those traits, only 2 neuroticism traits and 9 GWAS datasets were found to have causal effects on musculoskeletal outcomes (ukb-b-4630, ebi-a-GCST005232, ukb-a-230, ieu-a-1007, ebi-a-GCST005327, ebi-a-GCST003770, ieu-b-4848, ebi-a-GCST002920, ieu-a-118). There were 28 significant causal effects exerted by the “neuroticism score” and 30 causal effects exerted by “neuroticism” traits (Figure 2, Supplementary Table 2). While both “neuroticism score” and “neuroticism” traits originate from different studies and datasets (ebi-a-GCST005327 and ieu-a-1007, respectively), they both are continuous variables reporting the score of the neuroticism domain. The effect sizes, measured as an influence exerted by a change of one standard deviation in the exposure, of the neuroticism score on musculoskeletal injuries ranged from -6.27 (exerted on peroneal tendinitis; SE = 2.49; OR = 0.002 (1.4×10^−5^ - 0.248)) to 1.05 (exerted on ankle and foot dislocations, sprains, or strains; SE = 0.41; OR = 2.9 (1.3 - 6.3)). Similarly, the greatest negative effect size for neuroticism was also on peroneal tendinitis (beta = -21.27, SE = 9.66, OR = 5.79×10^−10^ (3.5×10^−18^ – 0.096)). The largest positive effect size for neuroticism was observed on dislocations, sprains and strains at the elbow (beta = 4.31, SE = 1.52, OR = 74.3 (3.8 – 1468). Overall, back pain appears to be the most frequently associated with a causal relationship to the neuroticism measures, appearing 15 times with neuroticism (50% of relationships) and 19 times with neuroticism score (68% of relationships).

**Figure 2:**
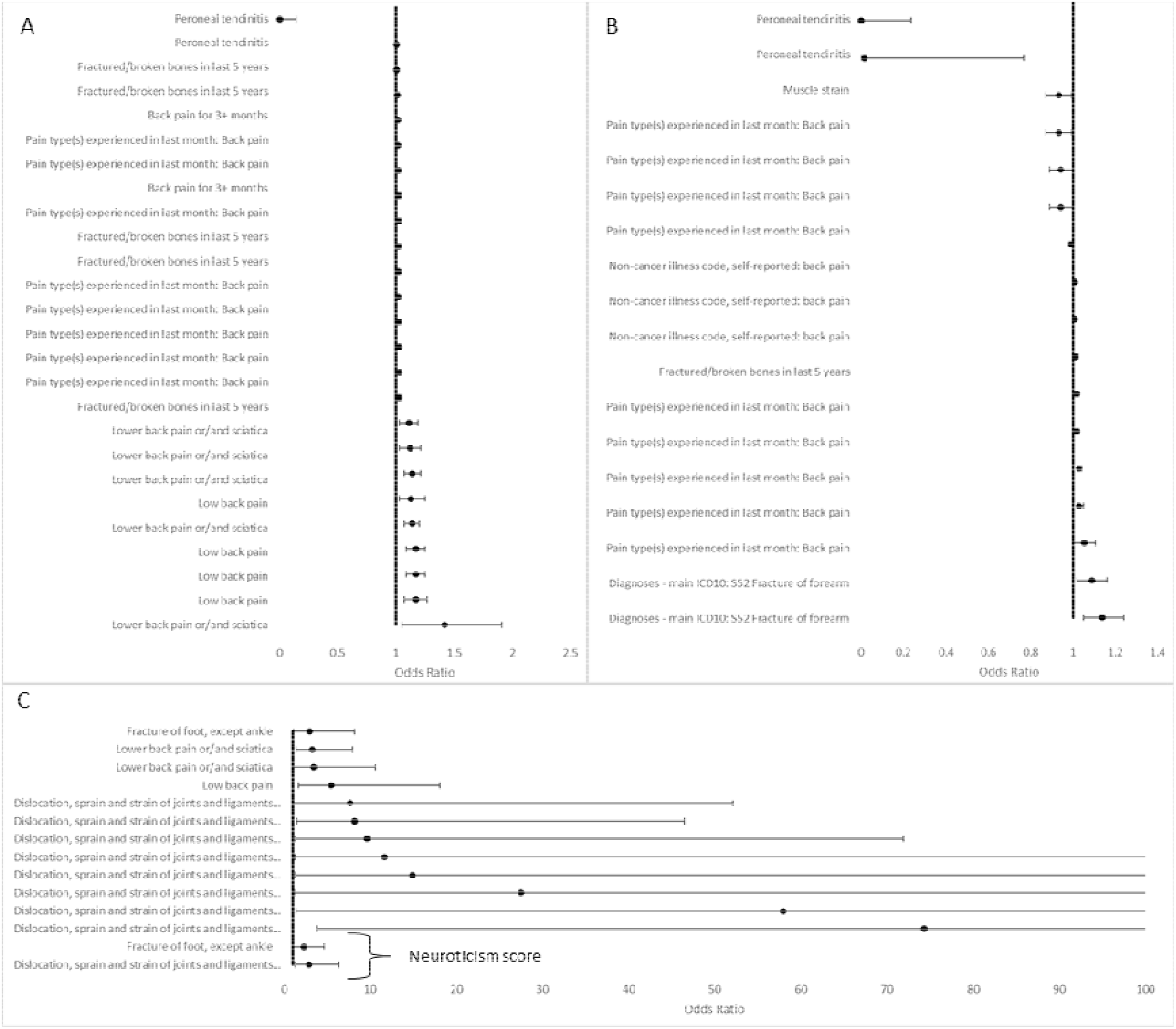
Odds ratios for “Big-5” domains with P < 5×10-8. A) Neuroticism score vs. MSI; B) Neuroticism vs. MSI; C) MR results with high odds ratios or wide confidence intervals for neuroticism (not indicated) or neuroticism score (indicated with bracket). All odds ratios are significant (P < 0.05) and the 95% confidence intervals do not overlap 1.0 (ratios provided as Supplementary Table 2).

We used the heterogeneity test provided within the 2SMR package to evaluate the degree of heterogeneity for each MR test. The package reported that there were enough variant instruments identified for 19 of the 58 MR tests (Supplemental Table 3). Using a four-group classification scheme based upon the I^2^ values [31, 32], we found 2 tests exhibited severe heterogeneity (I^2^ > 50%), 11 exhibited moderate heterogeneity (25% < I^2^ < 50%), and 3 each had mild or no heterogeneity (I^2^ < 25% or 0%, respectively). The severe heterogeneity appeared in the MR tests for neuroticism vs. lower back pain and forearm fractures calculated with the IVW estimator (I^2^ = 71% and 60%, and *P* = 0.001 and 0.02, respectively). Those with mild or no heterogeneity with neuroticism as the exposure were elbow injuries and foot fractures and were both calculated with the IVW estimator (I^2^ = 0% for both, *P* = 0.66 and 0.99, respectively). The neuroticism score exposure’s influence on lower back pain calculated using the Egger regression and on low back pain, foot fractures, and peroneal tendinitis calculated with the IVW estimator also had mild or no heterogeneity (I^2^ = 24%, 20%, 9%, and 0%, respectively; *P* = 0.05, 0.09, 0.28, and 0.51, respectively). In addition to heterogeneity, we observed some small, non-zero directional pleiotropy in 3 neuroticism estimates and 4 neuroticism score estimates (Table 4).

**Table 4:** Pleiotropy tests for causal relationships of personality on MSI

| Exposure<br>(IEU OpenGWAS ID) | Outcome<br>(IEU OpenGWAS ID) | Egger<br>Intercept | Lower<br>95% CI | Upper<br>95% CI | P-value |
| --- | --- | --- | --- | --- | --- |
| Neuroticism<br>(ebi-a-GCST005232) | Non-cancer illness code, self-reported: back pain<br>(ukb-b-11241) | 0.00032 | 0.00004 | 0.00059 | 0.028 |
| Neuroticism<br>(ebi-a-GCST005327) | Diagnoses - main ICD10: S52<br>Fracture of forearm<br>(ukb-a-590) | -0.0020 | -0.0033 | -0.0007 | 0.032 |
| Neuroticism (ieu-a-1007) | Diagnoses - main ICD10: S52<br>Fracture of forearm<br>(ukb-a-590) | -0.0019 | -0.0033 | -0.0004 | 0.042 |
| Neuroticism score<br>(ukb-a-230) | Fracture of foot, except ankle<br>(finn-b-ST19_FRACT_FOOT_ANKLE) | -0.0482 | -0.0896 | -0.0069 | 0.026 |
| Neuroticism score<br>(ukb-a-230) | Fractured/broken bones in last 5<br>years (ukb-a-308) | -0.0015 | -0.0027 | -0.0003 | 0.017 |
| Neuroticism score<br>(ukb-a-230) | Fractured/broken bones in last 5<br>years (ukb-b-13346) | -0.0012 | -0.0022 | -0.0001 | 0.036 |
| Neuroticism score<br>(ukb-b-4630) | Fractured/broken bones in last 5<br>years (ukb-a-308) | -0.0009 | -0.0018 | -0.0001 | 0.039 |

Relaxing the threshold for extracting instrumental variables to 5×10^−5^, we found 1,696 distinct SNPs across 14 personality GWAS datasets to use as instrumental variables compared to 210 GWAS significant SNPs in 8 personality GWAS datasets, an 8-fold increase in the instrumental variables from an almost doubling of the available datasets. Performing MR with these variables resulted in identifying nearly 100 more potential causal effects (Supplementary Table 3), and 119 estimates resulted in odds ratios not overlapping 1 (Supplementary Figure 1). There were 37 significant causal effects exerted by the neuroticism score, 86 by neuroticism, and a single causal effect exerted by conscientiousness on elbow injuries (Supplementary Figure 1, Supplementary Table 3).

### The causal influence of the aspects of emotional and social functioning on resilience to injury

From the 16 psychological traits associated with successfully completing CCT training that were obtained from the EQI and were unrelated to personality, we found 20 GWAS datasets identified related to those traits and 12 with causal effects on musculoskeletal outcomes (Figure 3, Supplementary Table 4). The largest positive direction effect size was for a diagnosis of anxiety on ankle or foot dislocations, sprains, and strains (beta = 6.0; SE = 2.6; OR = 410.2 (95% CI = 2.5 – 68371)) while the greatest negative direction effect size was for a visit to a general practice physician for anxiety on general muscle strains (beta = -16.8; SE = 8.1; OR = 5×10^−8^ (95% CI = 6.82×10^=15^ – 0.37). As with the personality traits, back pain appears to be most frequently related as an outcome to psychological traits, occurring in 4 of the relationships.

**Figure 3:**
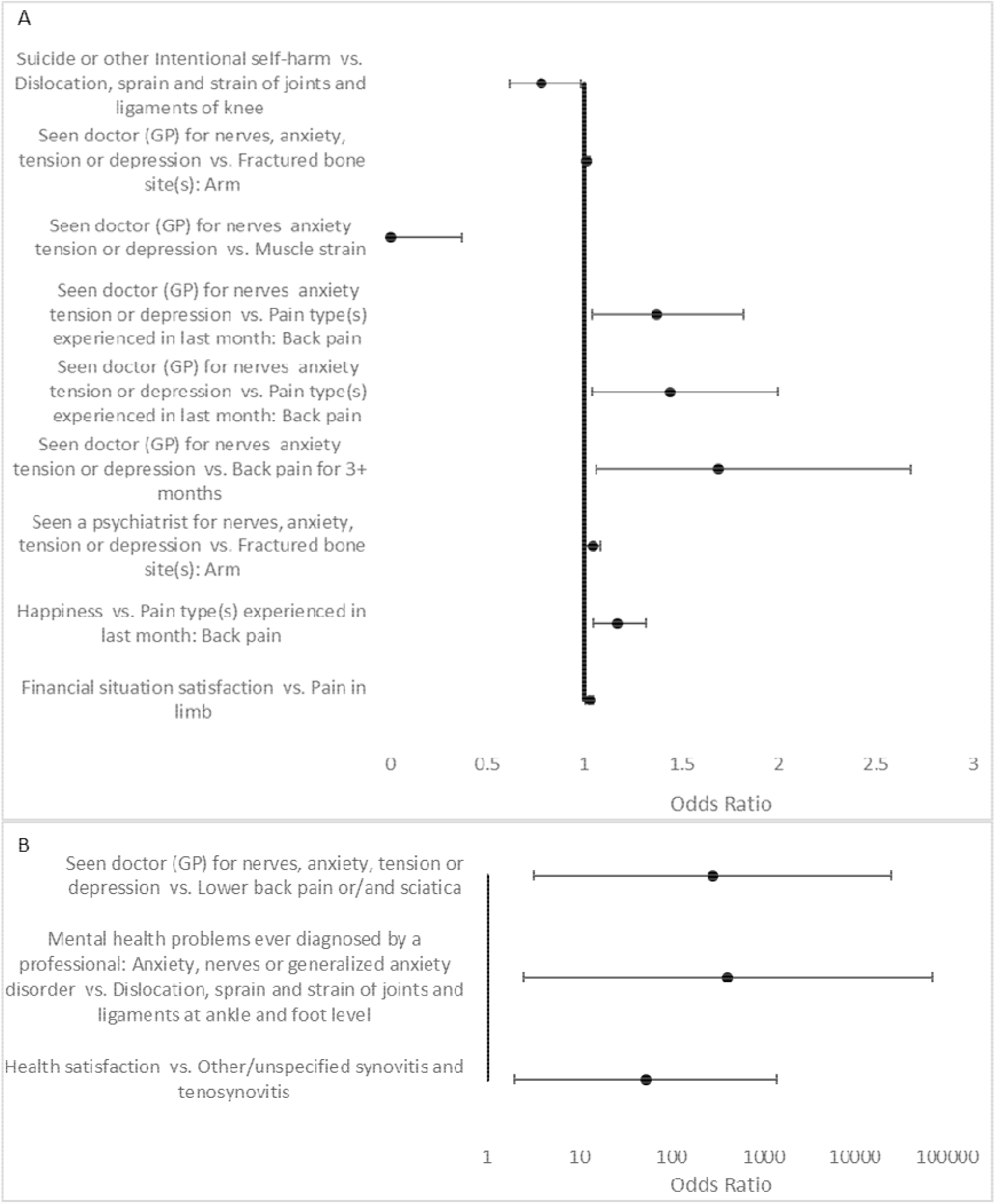
Odds ratios for instruments associated with non-“Big 5” personality domains extracted at *P* < 5×10^−8^. The causal trait is listed alongside brackets enclosing the affected musculoskeletal injuries. All odds ratios are significant (*P* < 0.05) and the 95% confidence intervals do not overlap 1.0 (ratios provided as Supplementary Table 4).

In our heterogeneity tests for the psychological traits that were not components of the “Big 5” personality domains, we observed enough variant instruments identified for 9 of the 17 MR tests (Supplemental Table 5). Using the same four-group I^2^ classification scheme as above [31, 32], we found no tests exhibited severe heterogeneity, 2 exhibited moderate heterogeneity, 1 had mild heterogeneity and the remaining 6 displayed no heterogeneity. The greatest degree of homogeneity was found for the effect of seeing a psychiatrist for nerves, anxiety, tension, or depression upon arm fractures (Q = 4.92, I^2^ = 39%, *P* = 0.18). With respect to pleiotropy, we only found a single relationship with non-zero pleiotropy: a history of having seen a general practice physician for nerves, anxiety, tension, or depression on lower leg or ankle fractures (Egger intercept = -0.0008, 95% CI = -0.0001 to -0.0015, *P* = 0.0357).

As with the personality-related assessments, when we relaxed the threshold for extracting instrumental variables to 5×10^−5^, we found an increase in the number of SNPs and GWAS datasets involved: 3,645 low threshold SNPs vs. 75 GWAS significant SNPs in 29 vs. 15 datasets, respectively. As expected, performing MR with the lower-threshold variables resulted in identifying more potential causal effects (2,344 vs. 375), and 98 estimates (vs. 58) with odds ratios not overlapping 1 (Supplementary Figure 2, Supplementary Table 6). Again, similar to the personality estimates, we found back pain to be the most frequently represented outcome with a causal relationship from psychological traits, appearing in 33 of the estimates with significant odds ratios.

## Discussion

In this report of our multiple parallel two-sample MR analyses, we found casual effects between neuroticism and several MSIs. This positive finding was confirmed by appearing in the results of multiple estimators (Wald ratio, inverse variance weighted, and weighted median analyses) and with various exposure and outcome GWAS summary statistics as input. However, no causal association were seen for the other “Big 5” personality domains (extraversion, conscientiousness, openness, or agreeableness). Furthermore, we also observed causal effects from psychological phenotypes associated with traits in successful CCT candidates on fractures and soft-tissue injuries, largely pain phenotypes. For instance, our results suggest that individuals with higher neuroticism scores will have an increased likelihood of experiencing lower back pain. In a recent study, while neuroticism was not significantly associated with debilitating lower back pain, high neuroticism was associated with use of coping strategies to control and adjust pain [33]. Both the recent results and our results support earlier evidence that higher neuroticism scores are related to increased pain catastrophizing and pain reports [34]. In fact, the relationship between personality domains and back pain has been researched dating at least back to 1993 [35], with neuroticism being found to mediate back pain perception throughout. While the effects of personality traits such as neuroticism on the pain threshold and catastrophizing are small, they may mediate the effects of occupational demands such as the physical loads and mental stresses encountered by CCT in the course of their jobs.

Past MR studies investigating causal effects on MSI have focused on physiological measurements. One two-sample MR study investigated the association of blood pressure on bone mineral density (BMD) and fractures using meta-analyses of both the exposure and outcome GWAS [36]. Interestingly, while the authors found a significant association between the blood pressure and forearm BMD, this effect did not extend to fractures or BMD in the femur or lumbar spine. Another study looked at the relationship between insulin-like growth factor-1 (IGF-1) levels on BMD and fractures [21]. In this case, the authors found that an increase in IGF-1 resulted in a 6% reduction of the odds of experiencing fractures, and after adjusting for the effect on BMD, that reduction was lower (4% reduction in odds) but still significant.

To the best of our knowledge, this is the first two-sample MR study to investigate and observe a causal relationship between psychology and MSI. The summary statistics we used for exposure and outcome phenotypes had very large sample sizes. Using two-sample MR with large datasets and multiple parallel sources, allows for us to observe any reproducible causal effects of psychological and personality traits in CCT on MSI risk, while at the same time minimizing reverse causation bias and confounding factors. In some of our results, we did observe some pleiotropy that may suggest other causal associations, especially for the effect of neuroticism on back pain, however the repeated observations of this interaction suggest that the observed effect is biologically meaningful.

Coupled together with our results, the prior reports of the impact of personality and psychology on MSI risk and recovery present a few interesting points to consider. As mentioned previously, the internal psychological traits of an individual have an impact on incurring injuries and also speeding recovery [6,13,15,16]. There are also, however, extrinsic factors that can influence these internal risk factors like skill and experience [37], instruction in broad-based coping skills [38], and even experiencing positive life events like weddings [10]. These observations of internal and external influences on psychology and MSI risk bring to mind the concepts of John Henryism and cynicism, and their potential roles in causing MSI. John Henryism, for instance, is the trait where an individual overcompensates for team members where they sacrifice themselves for the good of the team, while a cynic would also think “I can do the task better.” Together, with consideration of our observations with neuroticism, these two personality traits may also influence MSI risk or recovery.

There are several limitations to our approach that must be considered when evaluating the causal relationships we report. In order for a genetic variant to be a valid IV, three assumptions must be met: the variant must be predictive of the exposure; the variant is independent of any confounders in the exposure-outcome association; and the variant only affects the outcome through the exposure, and not through confounders or the outcome directly [39], It is likely that some of the genetic variants identified do not satisfy the IV assumptions because we used such large-scale GWAS datasets. Therefore, we included an analysis of the heterogeneity and found that, indeed, some of the causal relationships displayed a significant amount of heterogeneity. In accordance with the guidance of Burgess [40], we conclude from the limitations of the heterogeneity in the data that the causal relationship between psychological traits and injury risk is biologically meaningful, however the strength of the association is still to be determined. Also in accordance with the guidance of Burgess regarding the use of summarized data and two-sample Mendelian randomization, we found that many of the GWAS datasets we used were from populations with similar ethnic origin, however we did not restrict our datasets based upon ethnicity, therefore some of the observed heterogeneity may arise from ethnicity-based allele frequency differences. A limitation also exists, therefore, in trying to generalize the results: while observations from MR using two samples with the same population structure would be more robust, those results may not be applicable in other populations; in contrast, with our study we included a wider range of populations, possibly introducing undue bias and complexity and increasing false discoveries. Nonetheless, the repeated observations of several causal relationships diminish the impact of these limitations and suggest a valid conclusion from our results.

In conclusion, we found that personality traits and psychological states do indeed affect the outcomes of musculoskeletal injury. We found that individuals with high neuroticism appear to be more resilient to peroneal tendinitis (inflammation of the tendons on the exterior of the foot), while individuals with low neuroticism appear to be more resilient to back pain and dislocations, sprains, and strains of the extremities. This is applicable to CCT in that successful CCT tend to have lower neuroticism scores than the civilian norm and also lower than those who enter, but do not complete, CCT training. Therefore, we expect that CCT may have an increased chance of not experiencing back pain or being able to tolerate higher levels of pain than civilians. There does not appear to be much causation influenced by extraversion, conscientiousness or agreeableness on the risk or resilience to musculoskeletal injuries and most injury types do not appear to be causally related to personality traits. Furthermore, we found that having visited a health care provider for anxiety, nerves, tension, or depression is related to an increase in the risk of fractures, back pain, and muscle strain. Overall, our results suggest that back pain is significantly affected by psychology, and this opens an opportunity for possible mitigation or non-pharmaceutical interventions.

## Data Availability

All data used for this study are available from genetics summary statistics databases.

https://gwas.mrcieu.ac.uk/

## Notes

### Competing Interest Statement

The authors have declared no competing interest.

### Funding Statement

This study did not receive any funding

