## Supplemental Materials for "Evaluating the Relationship between Psychological Traits and Resilience to Musculoskeletal Injuries in Combat Control Graduates by Mendelian Randomization"

**Supplemental Table 1:** IEU OpenGWAS Project study identifiers for musculoskeletal injury phenotypes used.

| ID | Trait | Year | Author | Consortium | PMID |
| --- | --- | --- | --- | --- | --- |
| ebi-a-GCST006980 | Fractures | 2019 | Morris | NA | 30598549 |
| finn-b-JOINTPAIN | Pain in joint | 2021 | NA | NA | NA |
| finn-b-M13_ACHILLESTEND | Achilles tendinitis | 2021 | NA | NA | NA |
| finn-b-M13_CALCIFICTEND | Calcific tendinitis of shoulder | 2021 | NA | NA | NA |
| finn-b-M13_CALCTENDINITIS | Calcific tendinitis | 2021 | NA | NA | NA |
| finn-b-M13_CHRONSYNOVITISHANDWRIST | Chronic crepitant synovitis/bursitis of hand and wrist/periarhritis of wrist | 2021 | NA | NA | NA |
| finn-b-M13_IMPINGEMENT | Impingement syndrome of shoulder | 2021 | NA | NA | NA |
| finn-b-M13_LOWBACKPAIN | Low back pain | 2021 | NA | NA | NA |
| finn-b-M13_LOWBACKPAINORANDSCIATICA | Lower back pain or/and sciatica | 2021 | NA | NA | NA |
| finn-b-M13_MUSCLESTRAIN | Muscle strain | 2021 | NA | NA | NA |
| finn-b-M13_PATELLARTEND | Patellar tendinitis | 2021 | NA | NA | NA |
| finn-b-M13_PATHODISLOCATIO | Pathological/recurrent dislocation and subluxation of joint, not elsewhere classified | 2021 | NA | NA | NA |
| finn-b-M13_PERONEALTEND | Peroneal tendinitis | 2021 | NA | NA | NA |
| finn-b-M13_RECUDISLOCATIO | Recurrent dislocation and subluxation of joint | 2021 | NA | NA | NA |
| finn-b-M13_ROTATORCUFF | Rotator cuff syndrome | 2021 | NA | NA | NA |
| finn-b-M13_SHOULDERBURSITIS | Bursitis of shoulder | 2021 | NA | NA | NA |
| finn-b-M13_TENDOSYNOVITISNAS | Other/unspecified synovitis and tenosynovitis | 2021 | NA | NA | NA |
| finn-b-M13_TRANSSYNOVITIS | Transient synovitis | 2021 | NA | NA | NA |
| finn-b-ST19_DISLO_SPRAIN_STRAIN_JOINTS_LIGAM_ANKLE_FOOT_LEVEL | Dislocation, sprain and strain of joints and ligaments at ankle and foot level | 2021 | NA | NA | NA |
| finn-b-ST19_DISLO_SPRAIN_STRAIN_JOINTS_LIGAM_ELBOW | Dislocation, sprain and strain of joints and ligaments of elbow | 2021 | NA | NA | NA |
| ID | Trait | Year | Author | Consortium | PMID |
| finn-b-ST19_DISLO_SPRAIN_STRAIN_JOINTS_LIGAM_KNEE | Dislocation, sprain and strain of joints and ligaments of knee | 2021 | NA | NA | NA |
| finn-b-ST19_DISLO_SPRAIN_STRAIN_JOINTS_LIGAM_THORAX | Dislocation, sprain and strain of joints and ligaments of thorax | 2021 | NA | NA | NA |
| finn-b-ST19_FRACT_FEMUR | Fracture of femur | 2021 | NA | NA | NA |
| finn-b-ST19_FRACT_FOOT_ANKLE | Fracture of foot, except ankle | 2021 | NA | NA | NA |
| finn-b-ST19_FRACT_FOREA | Fracture of forearm | 2021 | NA | NA | NA |
| finn-b-ST19_FRACT_LUMBAR_SPINE_PELVIS | Fracture of lumbar spine and pelvis | 2021 | NA | NA | NA |
| finn-b-ST19_FRACT_NECK | Fracture of neck | 2021 | NA | NA | NA |
| finn-b-ST19_FRACT_RIBS_STERNUM_THORACIC_SPINE | Fracture of rib(s), sternum and thoracic spine | 2021 | NA | NA | NA |
| finn-b-ST19_FRACT_SHOUL_UPPER_ARM | Fracture of shoulder and upper arm | 2021 | NA | NA | NA |
| finn-b-ST19_FRACT_WRIST_HAND_LEVEL | Fracture at wrist and hand level | 2021 | NA | NA | NA |
| finn-b-ST19_INJURI_SHOUL_UPPER_ARM | Injuries to the shoulder and upper arm | 2021 | NA | NA | NA |
| ukb-a-308 | Fractured/broken bones in last 5 years | 2017 | Neale | Neale Lab | NA |
| ukb-a-341 | Neck/shoulder pain for 3+ months | 2017 | Neale | Neale Lab | NA |
| ukb-a-346 | Back pain for 3+ months | 2017 | Neale | Neale Lab | NA |
| ukb-a-353 | Knee pain for 3+ months | 2017 | Neale | Neale Lab | NA |
| ukb-a-381 | Leg pain on walking | 2017 | Neale | Neale Lab | NA |
| ukb-a-438 | Fractured bone site(s): Ankle | 2017 | Neale | Neale Lab | NA |
| ukb-a-439 | Fractured bone site(s): Wrist | 2017 | Neale | Neale Lab | NA |
| ukb-a-440 | Fractured bone site(s): Arm | 2017 | Neale | Neale Lab | NA |
| ukb-a-441 | Fractured bone site(s): Other bones | 2017 | Neale | Neale Lab | NA |
| ukb-a-472 | Pain type(s) experienced in last month: Neck or shoulder pain | 2017 | Neale | Neale Lab | NA |
| ID | Trait | Year | Author | Consortium | PMID |
| ukb-a-473 | Pain type(s) experienced in last month: Back pain | 2017 | Neale | Neale Lab | NA |
| ukb-a-475 | Pain type(s) experienced in last month: Hip pain | 2017 | Neale | Neale Lab | NA |
| ukb-a-476 | Pain type(s) experienced in last month: Knee pain | 2017 | Neale | Neale Lab | NA |
| ukb-a-590 | Diagnoses - main ICD10: S52 Fracture of forearm | 2017 | Neale | Neale Lab | NA |
| ukb-b-10387 | Leg pain on walking | 2018 | Ben Elsworth | MRC-IEU | NA |
| ukb-b-10873 | Diagnoses - main ICD10: M54.59 Low back pain (Site unspecified) | 2018 | Ben Elsworth | MRC-IEU | NA |
| ukb-b-11241 | Non-cancer illness code, self-reported: back pain | 2018 | Ben Elsworth | MRC-IEU | NA |
| ukb-b-12864 | Leg pain when walking normally | 2018 | Ben Elsworth | MRC-IEU | NA |
| ukb-b-13019 | Diagnoses - main ICD10: M25.5 Pain in joint | 2018 | Ben Elsworth | MRC-IEU | NA |
| ukb-b-133 | Hip pain for 3+ months | 2018 | Ben Elsworth | MRC-IEU | NA |
| ukb-b-13346 | Fractured/broken bones in last 5 years | 2018 | Ben Elsworth | MRC-IEU | NA |
| ukb-b-13979 | Non-cancer illness code, self-reported: fracture lower leg / ankle | 2018 | Ben Elsworth | MRC-IEU | NA |
| ukb-b-1418 | Diagnoses - main ICD10: M75.4 Impingement syndrome of shoulder | 2018 | Ben Elsworth | MRC-IEU | NA |
| ukb-b-1557 | Diagnoses - main ICD10: M54.5 Low back pain | 2018 | Ben Elsworth | MRC-IEU | NA |
| ukb-b-15582 | Fractured bone site(s): Ankle | 2018 | Ben Elsworth | MRC-IEU | NA |
| ukb-b-16118 | Neck/shoulder pain for 3+ months | 2018 | Ben Elsworth | MRC-IEU | NA |
| ID | Trait | Year | Author | Consortium | PMID |
| ukb-b-16254 | Pain type(s) experienced in last month: Knee pain | 2018 | Ben Elsworth | MRC-IEU | NA |
| ukb-b-17738 | Fractured bone site(s): Other bones | 2018 | Ben Elsworth | MRC-IEU | NA |
| ukb-b-18389 | Fractured heel | 2018 | Ben Elsworth | MRC-IEU | NA |
| ukb-b-18596 | Pain type(s) experienced in last month: Neck or shoulder pain | 2018 | Ben Elsworth | MRC-IEU | NA |
| ukb-b-19255 | Fractured bone site(s): Arm | 2018 | Ben Elsworth | MRC-IEU | NA |
| ukb-b-19813 | Diagnoses - main ICD10: M54.56 Low back pain (Lumbar region) | 2018 | Ben Elsworth | MRC-IEU | NA |
| ukb-b-2083 | Diagnoses - main ICD10: M25.56 Pain in joint (Lower leg) | 2018 | Ben Elsworth | MRC-IEU | NA |
| ukb-b-2804 | Diagnoses - main ICD10: S52.50 Fracture of lower end of radius (closed) | 2018 | Ben Elsworth | MRC-IEU | NA |
| ukb-b-3196 | Leg pain when walking uphill or hurrying | 2018 | Ben Elsworth | MRC-IEU | NA |
| ukb-b-3499 | Fractured heel (right) | 2018 | Ben Elsworth | MRC-IEU | NA |
| ukb-b-3665 | Leg pain on walking : effect of standing still | 2018 | Ben Elsworth | MRC-IEU | NA |
| ukb-b-3798 | Fractured bone site(s): Leg | 2018 | Ben Elsworth | MRC-IEU | NA |
| ukb-b-4122 | Non-cancer illness code, self-reported: joint pain | 2018 | Ben Elsworth | MRC-IEU | NA |
| ukb-b-4361 | Leg pain when standing still or sitting | 2018 | Ben Elsworth | MRC-IEU | NA |
| ukb-b-4921 | Diagnoses - main ICD10: M75.0 Adhesive capsulitis of shoulder | 2018 | Ben Elsworth | MRC-IEU | NA |
| ukb-b-50 | Diagnoses - main ICD10: M75.1 Rotator cuff syndrome | 2018 | Ben Elsworth | MRC-IEU | NA |
| ID | Trait | Year | Author | Consortium | PMID |
| ukb-b-5946 | Diagnoses - main ICD10: S82.80 Fractures of other parts of lower leg (closed) | 2018 | Ben Elsworth | MRC-IEU | NA |
| ukb-b-7118 | Diagnoses - main ICD10: M23.23 Derangement of meniscus due to old tear or injury (Medial collateral ligament or Other and unspecified medial meniscus) | 2018 | Ben Elsworth | MRC-IEU | NA |
| ukb-b-7289 | Pain type(s) experienced in last month: Hip pain | 2018 | Ben Elsworth | MRC-IEU | NA |
| ukb-b-7831 | Non-cancer illness code, self-reported: fracture wrist / colles fracture | 2018 | Ben Elsworth | MRC-IEU | NA |
| ukb-b-8213 | Diagnoses - main ICD10: M23.2 Derangement of meniscus due to old tear or injury | 2018 | Ben Elsworth | MRC-IEU | NA |
| ukb-b-8221 | Fractured heel (left) | 2018 | Ben Elsworth | MRC-IEU | NA |
| ukb-b-8463 | Back pain for 3+ months | 2018 | Ben Elsworth | MRC-IEU | NA |
| ukb-b-873 | Fractured bone site(s): Spine | 2018 | Ben Elsworth | MRC-IEU | NA |
| ukb-b-8906 | Knee pain for 3+ months | 2018 | Ben Elsworth | MRC-IEU | NA |
| ukb-b-9054 | Leg pain in calf/calves | 2018 | Ben Elsworth | MRC-IEU | NA |
| ukb-b-9571 | Fractured bone site(s): Wrist | 2018 | Ben Elsworth | MRC-IEU | NA |
| ukb-b-9694 | Diagnoses - main ICD10: M23.22 Derangement of meniscus due to old tear or injury (Posterior cruciate ligament or Posterior horn of medial meniscus) | 2018 | Ben Elsworth | MRC-IEU | NA |
| ID | Trait | Year | Author | Consortium | PMID |
| ukb-b-9708 | Diagnoses - main ICD10: M79.66 Pain in limb (Lower leg) | 2018 | Ben Elsworth | MRC-IEU | NA |
| ukb-b-9838 | Pain type(s) experienced in last month: Back pain | 2018 | Ben Elsworth | MRC-IEU | NA |
| ukb-d-M13_ADHCAPSULITIS | Adhesive capsulitis of shoulder | 2018 | Neale lab | NA | NA |
| ukb-d-M13_IMPINGEMENT | Impingement syndrome of shoulder | 2018 | Neale lab | NA | NA |
| ukb-d-M13_LIMBPAIN | Pain in limb | 2018 | Neale lab | NA | NA |
| ukb-d-M13_LOWBACKPAIN | Low back pain | 2018 | Neale lab | NA | NA |
| ukb-d-M13_ROTATORCUFF | Rotator cuff syndrome | 2018 | Neale lab | NA | NA |
| ukb-d-M65 | Diagnoses - main ICD10: M65 Synovitis and tenosynovitis | 2018 | Neale lab | NA | NA |
| ukb-d-S62 | Diagnoses - main ICD10: S62 Fracture at wrist and hand level | 2018 | Neale lab | NA | NA |
| ukb-d-S72 | Diagnoses - main ICD10: S72 Fracture of femur | 2018 | Neale lab | NA | NA |
| ukb-d-S82 | Diagnoses - main ICD10: S82 Fracture of lower leg, including ankle | 2018 | Neale lab | NA | NA |

**Supplemental Table 2:** NEO personality domain MR results with instrumental variables extracted at *P* < 5x10^-8^

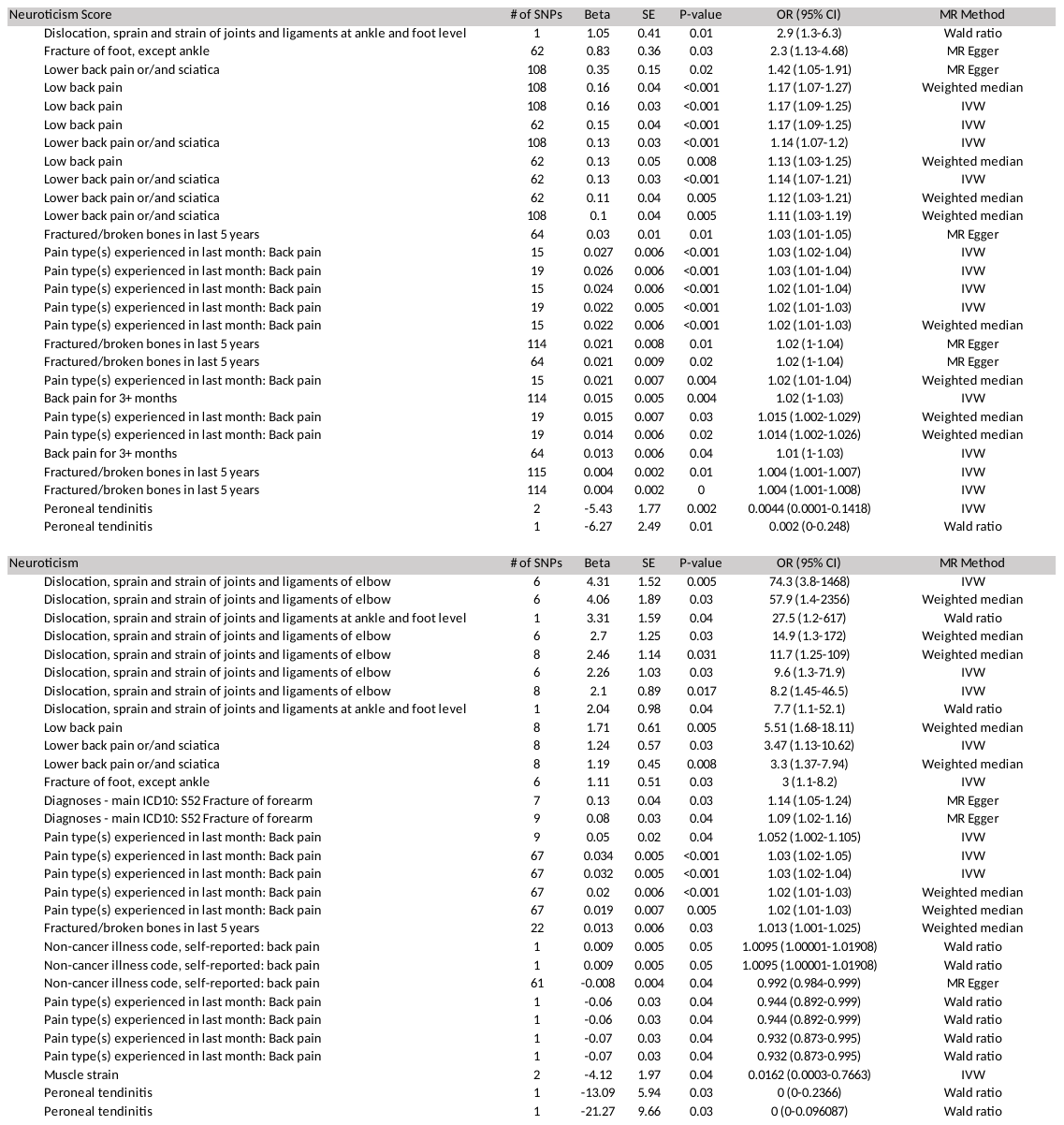

**Supplemental Table 3:** Heterogeneity analysis for personality-related instruments extracted at *P*<5x10^-8^.

| Exposure | Outcome | MR Method | Q | df | I^2^ | *P*-value |
| --- | --- | --- | --- | --- | --- | --- |
| Neuroticism | Lower back pain or/and sciatica | IVW | 23.78 | 7 | 71% | 0.001 |
| Neuroticism | Diagnoses - main ICD10: S52 Fracture of forearm | IVW | 15.14 | 6 | 60% | 0.02 |
| Neuroticism | Pain type(s) experienced in last month: Back pain | IVW | 121.91 | 66 | 46% | <0.001 |
| Neuroticism | Pain type(s) experienced in last month: Back pain | IVW | 116.87 | 66 | 44% | <0.001 |
| Neuroticism score | Pain type(s) experienced in last month: Back pain | IVW | 31.55 | 18 | 43% | 0.03 |
| Neuroticism score | Fractured/broken bones in last 5 years | IVW | 96.95 | 63 | 35% | 0.004 |
| Neuroticism score | Pain type(s) experienced in last month: Back pain | MR Egger | 19.45 | 13 | 33% | 0.11 |
| Neuroticism score | Fractured/broken bones in last 5 years | IVW | 165.76 | 113 | 32% | <0.001 |
| Neuroticism score | Back pain for 3+ months | MR Egger | 90.13 | 62 | 31% | 0.01 |
| Neuroticism score | Lower back pain or/and sciatica | MR Egger | 151.76 | 106 | 30% | 0.002 |
| Neuroticism score | Low back pain | IVW | 151.53 | 107 | 29% | 0.003 |
| Neuroticism score | Pain type(s) experienced in last month: Back pain | IVW | 19.61 | 14 | 29% | 0.14 |
| Neuroticism score | Fractured/broken bones in last 5 years | MR Egger | 83.28 | 62 | 26% | 0.04 |
| Neuroticism score | Lower back pain or/and sciatica | MR Egger | 78.77 | 60 | 24% | 0.05 |
| Neuroticism score | Low back pain | IVW | 76.32 | 61 | 20% | 0.09 |
| Neuroticism score | Fracture of foot, except ankle | IVW | 66.93 | 61 | 9% | 0.28 |
| Neuroticism | Dislocation, sprain and strain of joints and ligaments of elbow | IVW | 3.27 | 5 | 0% | 0.66 |
| Neuroticism | Fracture of foot, except ankle | IVW | 0.57 | 5 | 0% | 0.99 |
| Neuroticism score | Peroneal tendinitis | IVW | 0.43 | 1 | 0% | 0.51 |

**Supplementary Figure 1**: Odds ratios for “Big 5” instruments with *P* < 5x10^-5^. The causal trait is listed alongside brackets enclosing the affected musculoskeletal injuries. All odds ratios are significant (*P* < 0.05) and the 95% confidence intervals do not overlap 1.0.

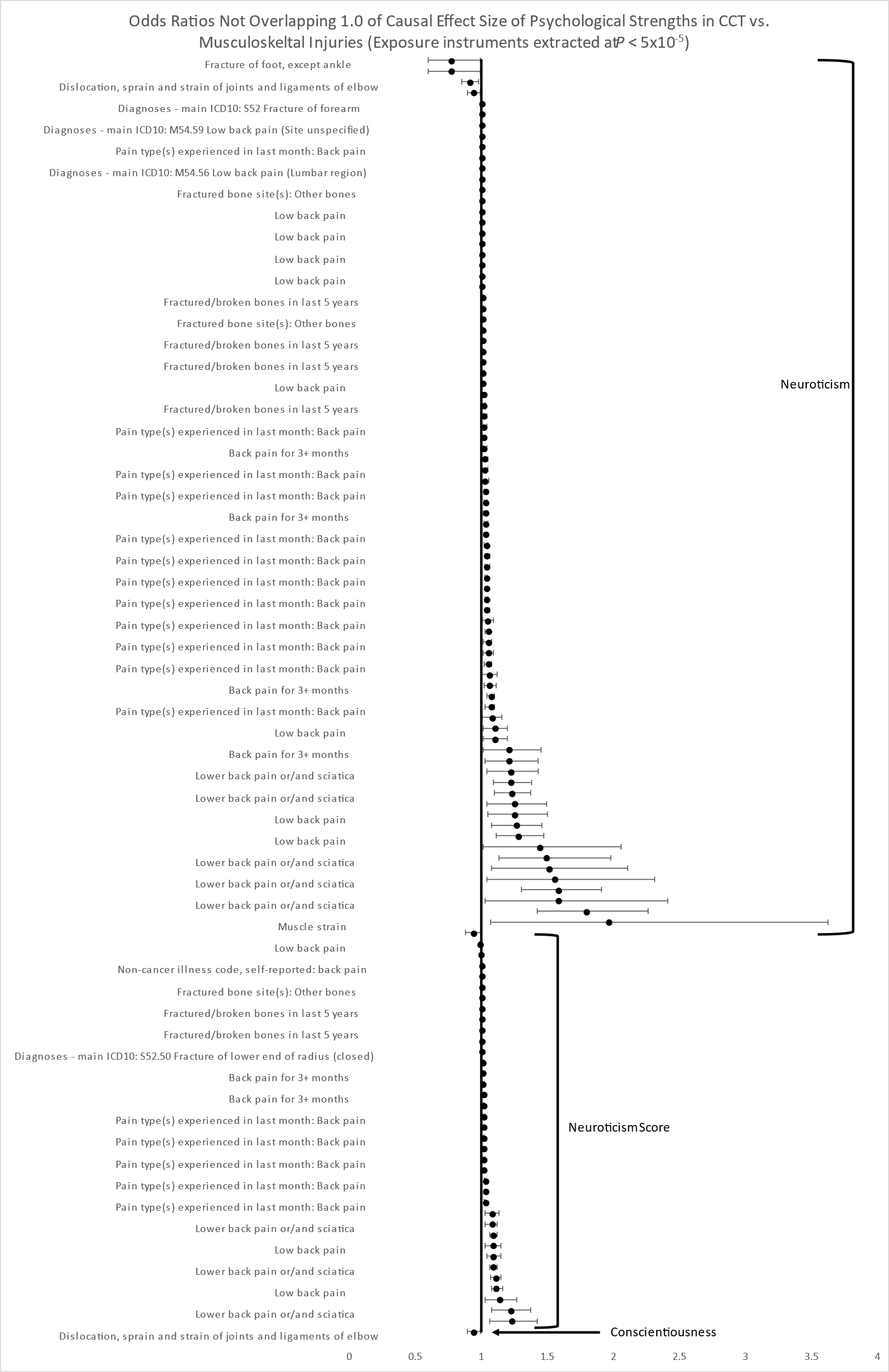

**Supplementary Table 3:** NEO personality domain MR results with instrumental variables extracted at *P* < 5x10^-5^

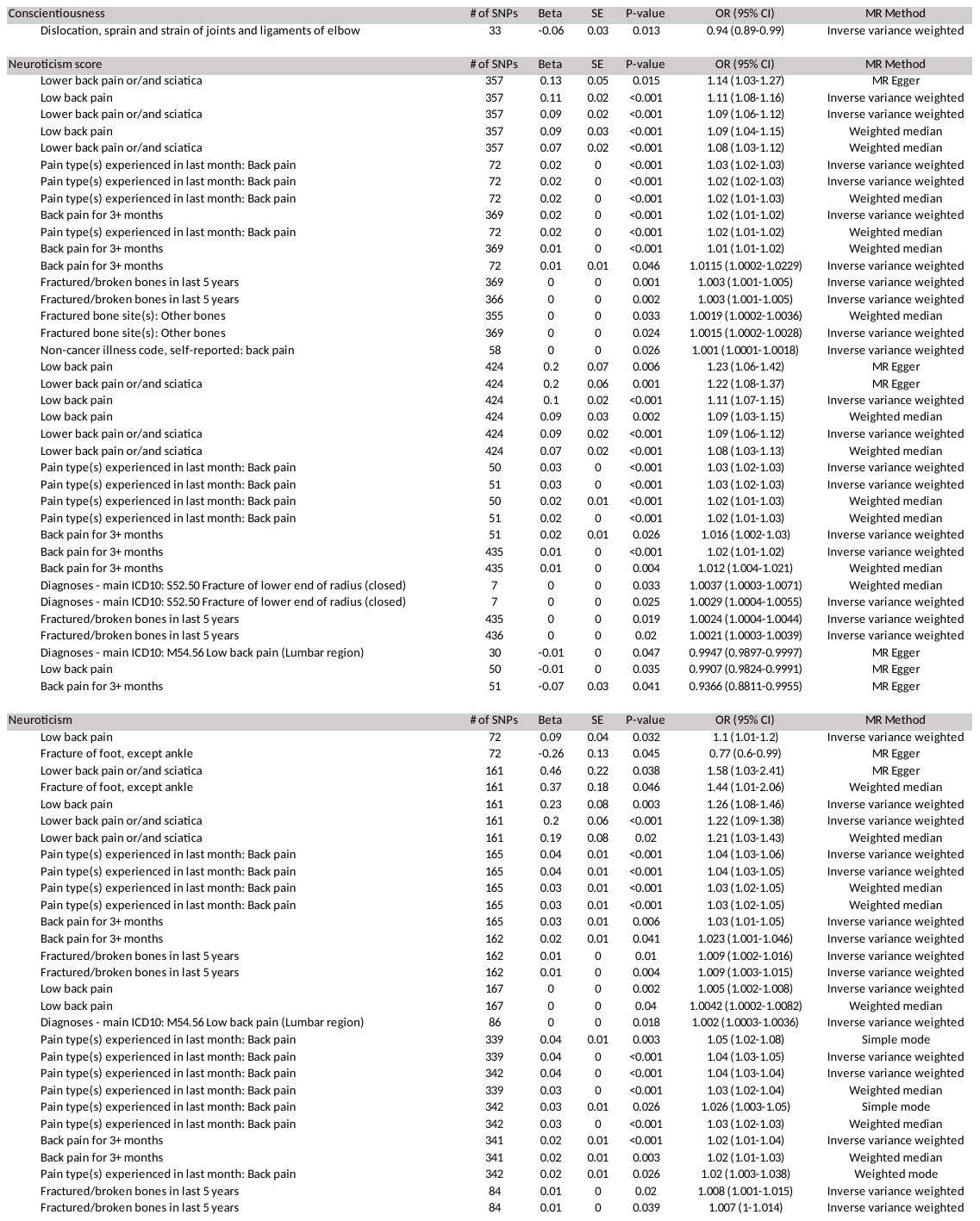

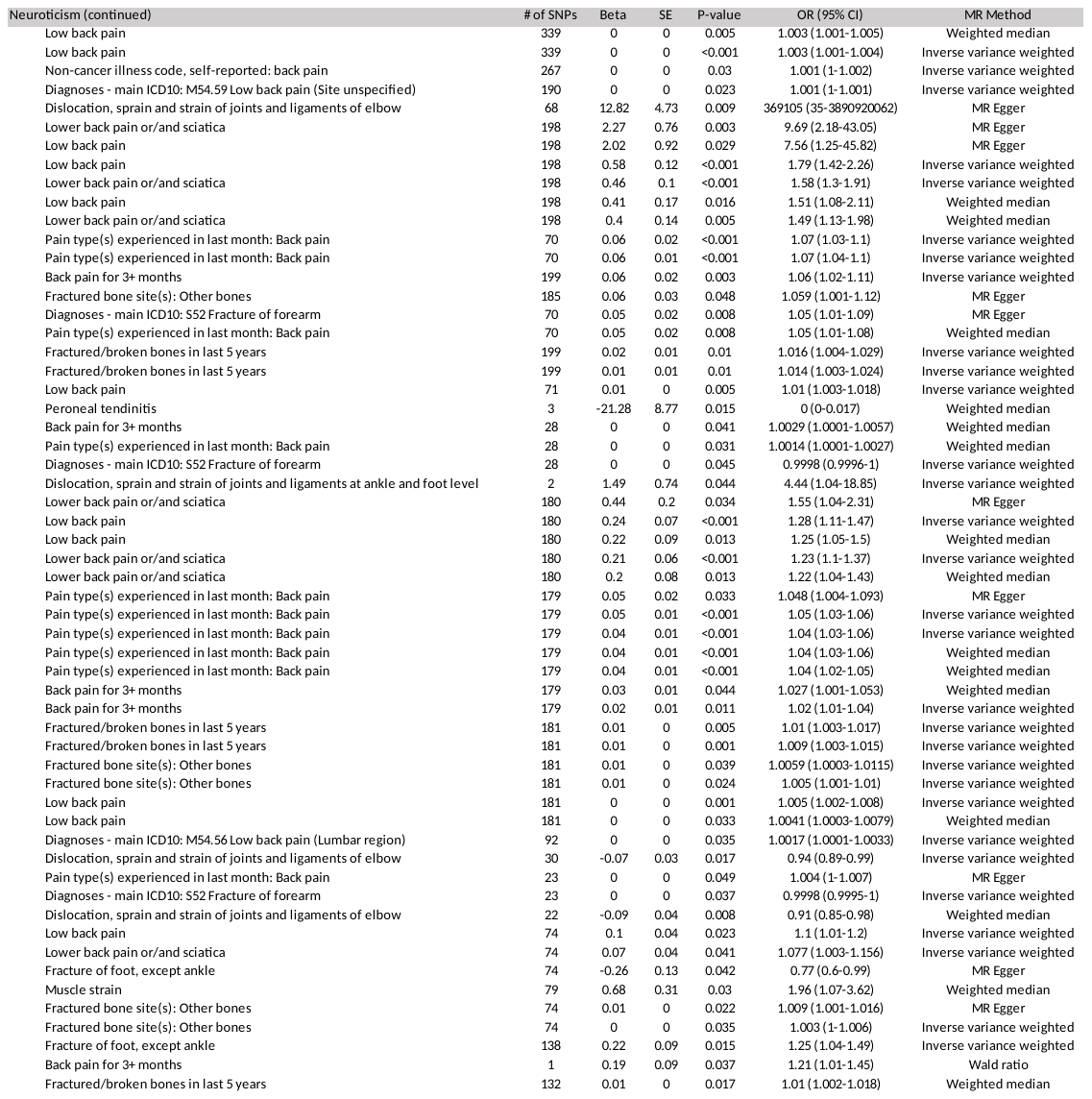

**Supplementary Table 4:** Non-personality trait MR results with instrumental variables extracted at *P* < 5x10^-8^

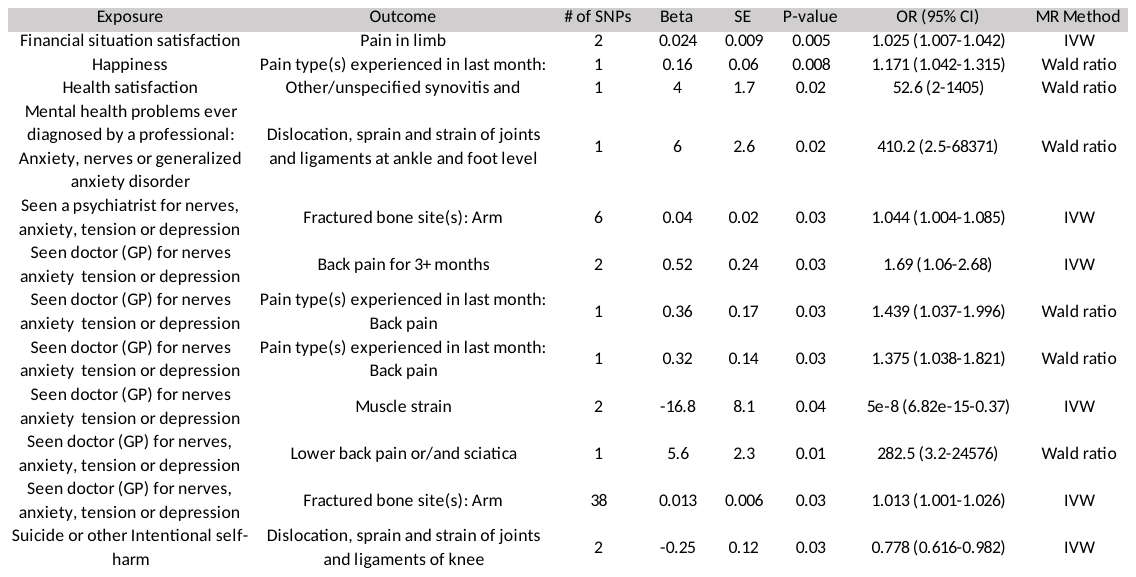

**Supplemental Table 5:** Heterogeneity analysis for psychological (non-personality related) instruments extracted at *P*<5x10^-8^.

| Exposure | Outcome | MR Method | Q | df | I^2^ | *P*-value |
| --- | --- | --- | --- | --- | --- | --- |
| Seen a psychiatrist for nerves, anxiety, tension or depression | Fractured bone site(s): Arm | IVW | 4.92 | 3 | 39% | 0.18 |
| Seen doctor (GP) for nerves, anxiety, tension or depression | Fractured bone site(s): Ankle | IVW | 52.6 | 39 | 26% | 0.07 |
| Seen doctor (GP) for nerves, anxiety, tension or depression | Fractured bone site(s): Arm | IVW | 46.0 | 37 | 20% | 0.15 |
| Suicide or other Intentional self-harm | Dislocation, sprain and strain of joints and ligaments of knee | IVW | 0.065 | 1 | 0% | 0.80 |
| Seen doctor (GP) for nerves anxiety tension or depression | Muscle strain | IVW | 0.017 | 1 | 0% | 0.90 |
| Seen doctor (GP) for nerves anxiety tension or depression | Back pain for 3+ months | IVW | 0.13 | 1 | 0% | 0.71 |
| Seen doctor (GP) for nerves anxiety tension or depression | Non-cancer illness code, self-reported: fracture lower leg / ankle | MR Egger | 6.99 | 12 | 0% | 0.86 |
| Seen a psychiatrist for nerves, anxiety, tension or depression | Fractured bone site(s): Arm | IVW | 3.31 | 5 | 0% | 0.65 |
| Financial situation satisfaction | Pain in limb | IVW | 0.80 | 1 | 0% | 0.37 |

MR= Mendelian randomization; IVW= Inverse variance weighted; GP= general practice

**Supplementary Figure 2**: Odds ratios for instruments associated with non-“Big 5” personality domains extracted at *P* < 5x10^-5^. The causal trait is listed alongside brackets enclosing the affected musculoskeletal injuries. All odds ratios are significant (*P* < 0.05) and the 95% confidence intervals do not overlap 1.0. A) Odds ratios less than 10 with narrow 95% confidence intervals; B) Odds ratios less than 10 with wide confidence intervals; C) Odds ratios greater than 10.

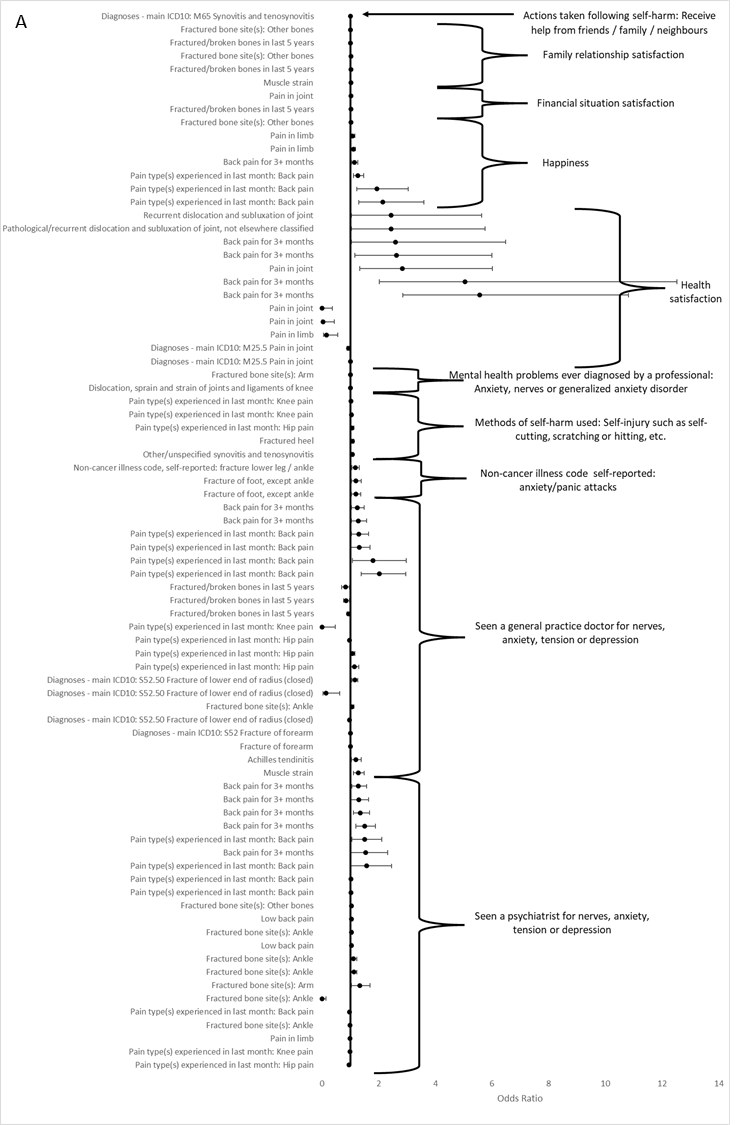

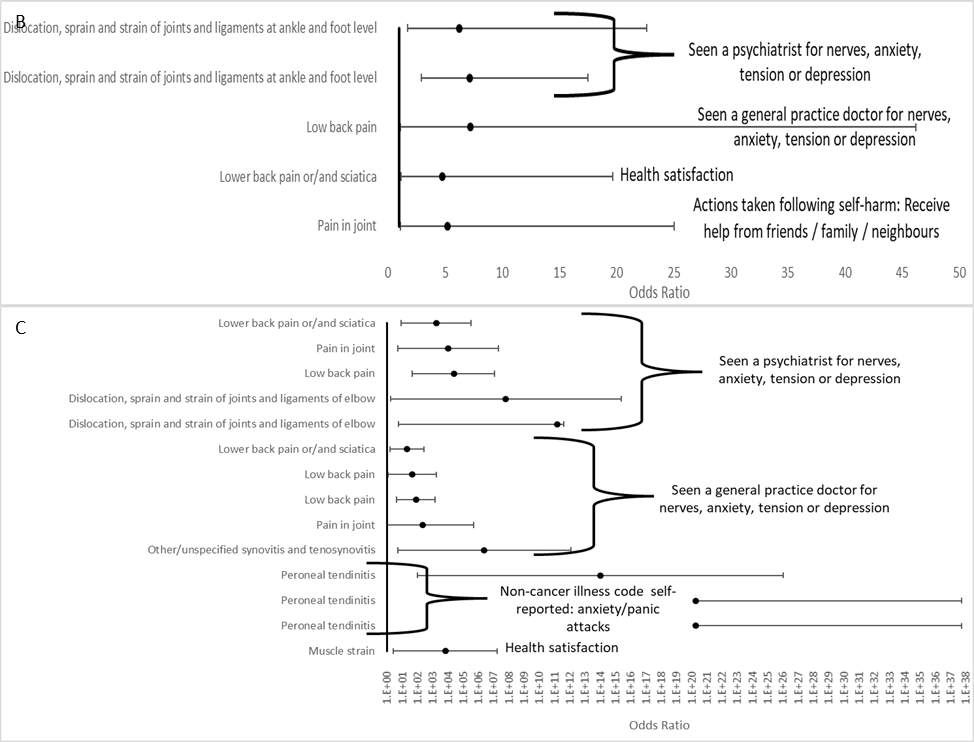

**Supplementary Table 5:** Non-personality trait MR results with instrumental variables extracted at *P* < 5x10^-5^

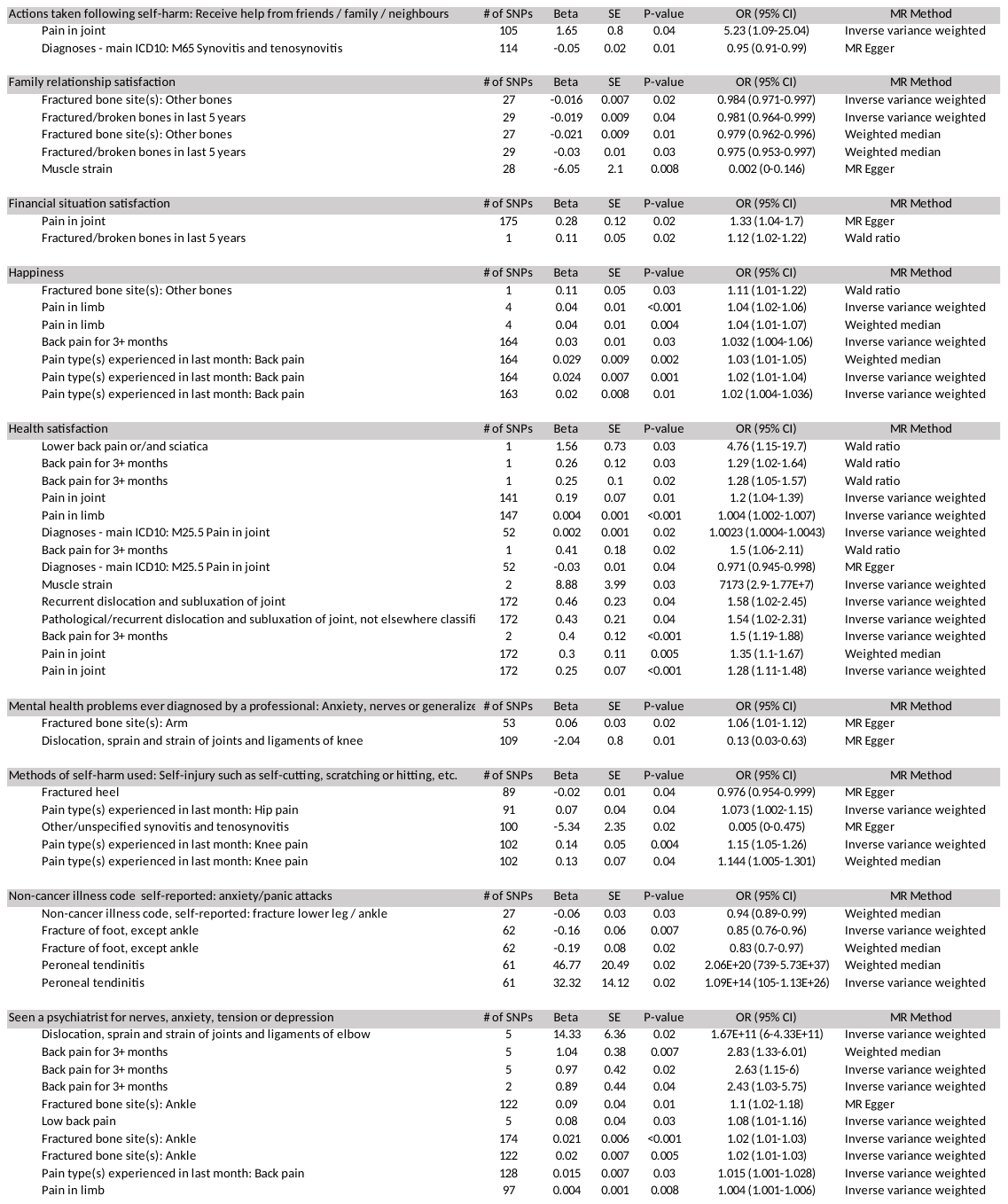

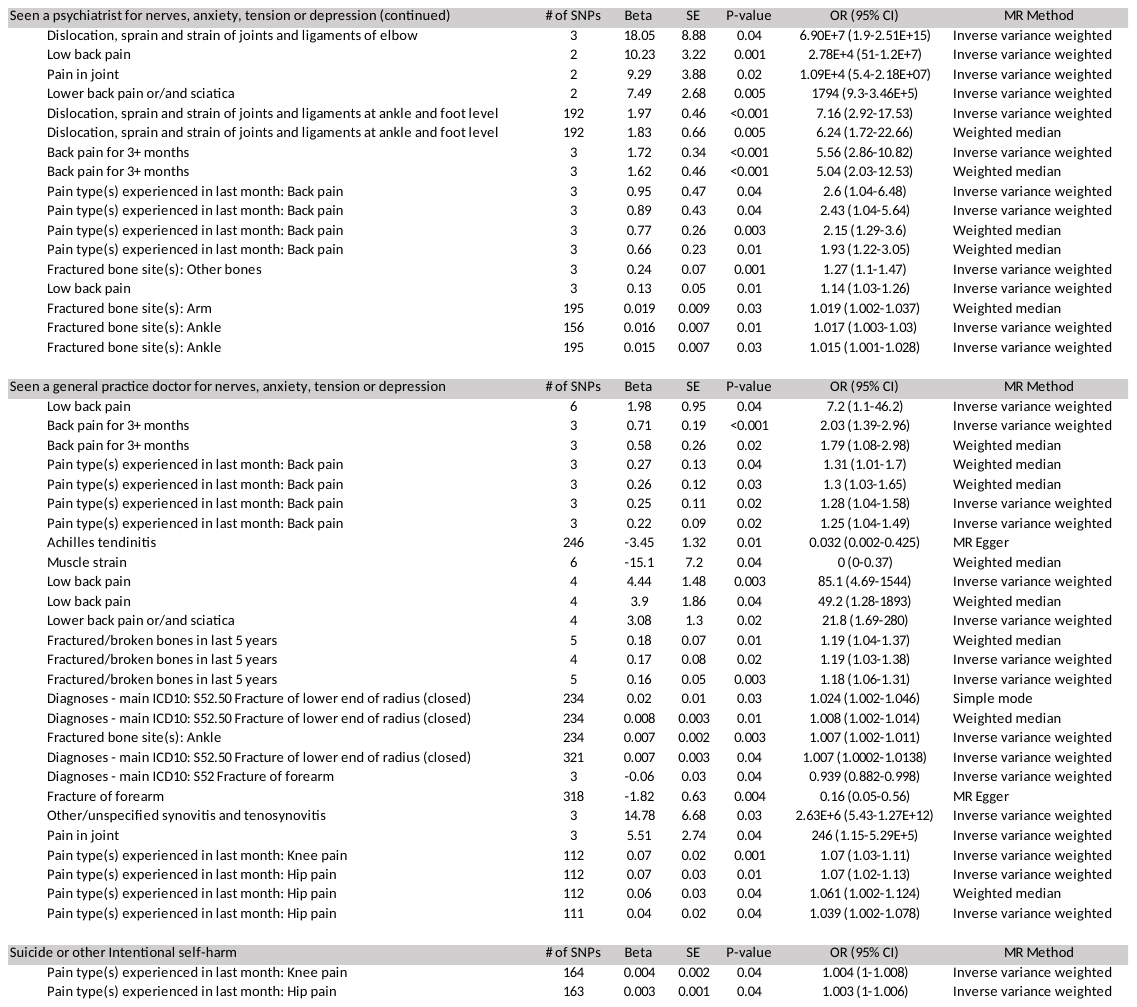
